# Heterologous prime-boost vaccination with ChAdOx1 nCoV-19 and BNT162b2 mRNA

**DOI:** 10.1101/2021.07.03.21258887

**Authors:** Matthias Tenbusch, Sofie Schumacher, Emanuel Vogel, Alina Priller, Jürgen Held, Philipp Steininger, Stephanie Beileke, Pascal Irrgang, Ronja Brockhoff, Jon Salmanton-García, Kathrin Tinnefeld, Hrvoje Mijocevic, Kilian Schober, Christian Bogdan, Sarah Yazici, Percy Knolle, Oliver A. Cornely, Klaus Überla, Ulrike Protzer

## Abstract

Administration of a first dose of the COVID-19 vaccine ChAdOx1 nCoV-19 (Vaxzevria^®^, AstraZeneca) is associated with a certain risk for vaccine-induced immune thrombotic thrombocytopenia. Therefore, several countries have recommended replacing the second dose of ChAdOx1 nCoV-19 with an mRNA-based vaccine as a precautionary measure, although data on safety and efficacy of such heterologous prime-boost regimen are sparse. Therefore, vaccinees, who had received a heterologous vaccination using ChAdOx1 nCoV-19 as prime and BNT162b2 (Comirnaty^®^, BioNTech-Pfizer) mRNA as boost vaccination were offered SARS-CoV-2 antibody testing to quantify their vaccine-induced neutralizing antibody response^5^. The results were compared to cohorts of healthcare workers or volunteers, who received homologous BNT162b2 or homologous ChAdOx1 nCoV-19 vaccination regimens, respectively. A striking increase of vaccine-induced SARS-CoV-2 neutralizing antibody activity was observed in 229 vaccinees that received a BNT162b2 boost 9 to 12 weeks after ChAdOx1 nCoV-19 prime. In our cohort comprising over 480 individuals, the heterologous vaccination scheme induced significantly higher neutralizing antibody titers than homologous ChAdOx1 nCoV-19 and even than homologous BNT162b2 vaccination. This proves that a single dose of a COVID-19 mRNA vaccine after ChAdOx1 nCoV-19 prime vaccination is sufficient to achieve high neutralizing antibody levels predicting immune protection from SARS-CoV-2 infection, and may even increase vaccine efficacy offering an alternative in a setting of vaccine shortage.

## Manuscript Text

Administration of a first dose of the COVID-19 vaccine ChAdOx1 nCoV-19 (Vaxzevria^®^, AstraZeneca) is associated with a risk for vaccine-induced immune thrombotic thrombocytopenia (VITT; also termed thrombosis thrombocytopenia syndrome, TTS) in the range of one to two cases per 100,000 vaccinations, with younger women showing the highest risk^1,2^. Fewer cases have been reported for the Ad26.COV2.S COVID-19 vaccine Janssen (Johnson & Johnson), another adenoviral-vector based COVID-19 vaccine^3^. As vaccine-induced antibodies against platelet factor 4 (PF4) have been implicated in the pathogenesis of VITT^1,2^, an amplifying effect of a second vaccine dose cannot be excluded. Therefore, several countries have recommended replacing the second dose of ChAdOx1 nCoV-19 with an mRNA-based vaccine as a precautionary measure, although data on safety and efficacy of such heterologous prime-boost regimen are sparse or even absent^4^.

Considering these uncertainties, vaccinees, who had received a heterologous vaccination using ChAdOx1 nCoV-19 as prime and BNT162b2 (Comirnaty^®^, BioNTech-Pfizer) mRNA as boost vaccination were offered SARS-CoV-2 antibody testing to quantify their vaccine-induced neutralizing antibody response^5^. The results were compared to cohorts of healthcare workers or volunteers, who received homologous BNT162b2 or homologous ChAdOx1 nCoV-19 vaccination regimens, respectively. Demographic data of the cohorts analyzed are presented in the Supplementary Appendix (Table S1 and S2).

To assess protective antibody responses, a surrogate neutralization assay (NAb assay, Yhlo, Shenzen, China) based on the competition of serum antibodies with recombinant angiotensin-converting-enzyme 2 for binding to the SARS-CoV-2 spike protein receptor-binding domain was performed in two certified, university virology diagnostic laboratories in Munich and Erlangen. Importantly, the surrogate neutralization assay correlated closely with a cell-culture based SARS-CoV-2 neutralization assay (Supplementary Appendix, Figure S1).

A striking increase of vaccine-induced SARS-CoV-2 neutralizing antibody activity was observed in 229 vaccinees that received a BNT162b2 boost 9 to 12 weeks after ChAdOx1 nCoV-19 prime. Sera were analyzed at the day of BNT162b2 boost vaccination and two weeks thereafter (Figure 1A). The single non-responder reported chronic lymphatic leukemia. High antibody levels already after ChAdOx1 nCoV-19 prime vaccination in two individuals most likely reflected prior, undetected SARS-CoV-2 infection.

**Fig. 1.**
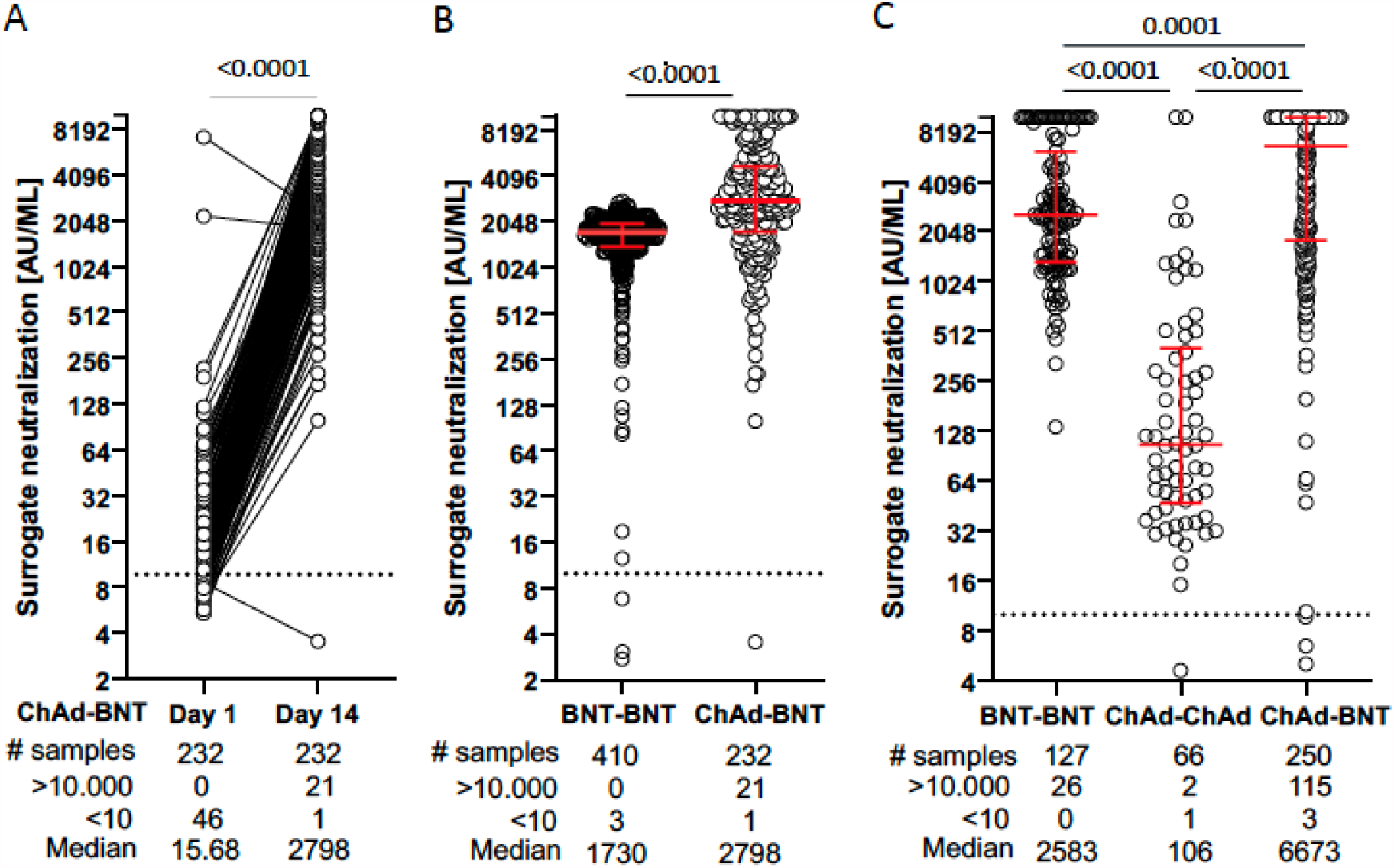
Comparison of surrogate neutralization activity induced by homologous and heterologous COVID-19 vaccine regimens. Vaccinees received either BNT162b2 mRNA (BNT) or ChAdOx1 nCoV-19 (ChAd) vaccine. (Panel A) In vaccinees who received ChAd as a prime and BNT as a boost vaccine, serum samples were obtained at the day of boost vaccination and at day 14±1 after the heterologous booster. Surrogate neutralization activity is given in paired samples. (Panel B) Dot plots showing surrogate neutralization activity from a group of vaccinees receiving homologous vaccination with BNT (BNT - BNT) compared to that receiving heterologous vaccination (ChAd – BNT) determined by laboratory 1. (Panel C) Dot plots of surrogate neutralization activity after homologous BNT - BNT, ChAd - ChAd and heterologous ChAd - BNT vaccination determined by laboratory 2. Dots represent single vaccinees. P values from a two-tailed Wilcoxon matched-pairs signed rank test (A), a two-tailed Mann Whitney test (B), and a Dunns multiple comparisons test (C) are shown above the graph. Median and interquartile ranges in (B) and (C) are indicated by red horizontal lines. Descriptive statistics shown below the graph include numbers (#) of samples, median and number of samples with results below the lower (<10) and above the upper (>10.000) cut-off of the surrogate neutralization assay.

Figures 1B and 1C show comparisons of the antibody levels observed two weeks after two doses of BNT162b2, two doses of ChAdOx1 nCoV-19 or one dose of ChAdOx1 nCoV-19 followed by one dose of BNT162b2. The heterologous vaccination induced significantly higher neutralizing antibody titers than homologous ChAdOx1 nCoV-19 and even than homologous BNT162b2 vaccination. Thus, a single dose of a COVID-19 mRNA vaccine after ChAdOx1 nCoV-19 prime vaccination is sufficient to achieve high neutralizing antibody levels predicting immune protection from SARS-CoV-2 infection^5^.

These results obtained in over 480 individuals, who received a prime with an adenoviral vector and a boost with an mRNA vaccine, indicate that a heterologous prime-boost with different COVID-19 vaccines may increase vaccine efficacy, and offers an alternative in a setting of vaccine shortage. Further studies, however, need to address the safety and clinical efficacy of heterologous vaccination regimens.

(479 words)

## Data Availability

We share data if reasonable requests are received. Requests should be directed to the corresponding author at

