## Supplemental Appendix for "Heterologous prime-boost vaccination with ChAdOx1 nCoV-19 and BNT162b2 mRNA"

### Study team (to be listed in PubMed)

University Hospital rechts der Isar, Technical University of Munich: Hedwig Roggendorf, Otto Zelger, Catharina Christa.
University Hospital of Cologne: Sarah Heringer.
University Hospital Erlangen: Rayya Alsalameh, Monika Wytopil.

### Table of contents:

1. Supplementary Table S1: Characteristics of cohorts analyzed in laboratory 1 (Munich, Germany)
2. Characteristics of cohorts analyzed in laboratory 1 (Munich, Germany)
3. Supplementary Table S2: Characteristics of cohorts analyzed in laboratory 2 (Erlangen, Germany)
4. Characteristics of cohorts analyzed in laboratory 2 (Erlangen, Germany)
5. Supplementary Figure S1: Correlation of the surrogate neutralizing antibody assay and a cell-culture based infection inhibition assay using SARS-CoV-2 wild-type
6. Ethics statement
7. Funding statement
8. Acknowledgements

1. **Table S1. Characteristics of cohorts analyzed in laboratory 1 (Munich, Germany)**

| | **ChAdOx1 nCoV-19 prime, BNT162b2 mRNA boost**<br>**CGN N = 183; MUC N = 49; total N = 232** | **BNT162b2 mRNA prime, BNT162b2 mRNA boost**<br>**MUC N = 410** |
|---|---|---|
| Volunteer source | General population at vaccination center | Healthcare workers |
| Age in years, median (IQR) [range] | 47 (33-55) [18-65] | 38 (31-48) [20-78] |
| Sex, n (%) Female | 190 (81.9%) | 248 (60.5%) |
| Sex, n (%) Male | 42 (18.1%) | 162 (39.5%) |
| Time from prime to boost dose in days, median (IQR) [range] | 63 (63-77) [54-85] | 21 (21-22) [18-25] |
| Time from boost dose to blood collection in days, median (IQR) [range] | 14 (13-15) [13-15] | 14 (13-15) [9-62] |

CGN, Cologne; MUC, Munich; IQR: interquartile range.

2. **Characteristics of cohorts analyzed in laboratory 1 (Munich, Germany)**

To evaluate standard mRNA vaccination, employees of the University Hospital rechts der Isar of the Technical University of Munich were offered to have their antibody responses to SARS-CoV-2 analyzed after vaccination with two doses of BNT162b2 mRNA. Persons with known or accidentally detected prior SARS-CoV-2 infection were excluded. Serum samples obtained from 410 persons at the time of booster vaccination and 14±1 day after the second vaccination were analyzed.

Given the uncertainties in the immunogenicity of heterologous prime boost vaccinations, 232 vaccinees, who received a first vaccination with ChAdOx1 nCoV-19 and - according to current recommendation in Germany - a booster vaccination with BNT162b2 mRNA at the vaccination center in Cologne or in Munich, Germany, were offered to register for blood sampling appointments at the day of booster vaccination with BNT162b2 mRNA and 13 to 15 days after the second vaccination. Information on age, sex, and dates of first and second vaccination were obtained. A summary of the characteristics of the different groups of vaccinees is shown in Table S1.

### 3. Table S2. Characteristics of cohorts analyzed by laboratory 2.

| | **BNT162b2 mRNA prime, BNT162b2 mRNA boost**<br><br>**ERL N = 127** | **ChAdOx1 nCoV-19 prime, ChAdOx1 nCoV-19 boost**<br><br>**ERL N = 66** | **ChAdOx1 nCoV-19 prime, BNT162b2 mRNA boost**<br><br>**ERL N = 250** |
|---|---|---|---|
| Volunteer source | Healthcare worker | General population at vaccination center | General population at vaccination center |
| Age in years, median (IQR) [range] | 41 (27-52) [20-65] | 57 (45-62) [31-64] | 52 (31-59) [19-59] |
| Sex, n (%) Female | 90 (70.87%) | 43 (65.2%) | 160 (64.0%) |
| Sex, n (%) Male | 37 (29.13%) | 23 (34.8%) | 90 (36.0%) |
| Time from prime to boost dose in days, median (IQR) [range] | 23 (22-25) [14-29] * | 63 (63-63) [63-63] | 63 (63-63) [57-71] |
| Time from boost dose to blood collection in days, median (IQR) [range] | 14 (14) [10-21] | 15 (13-15) [13-16] | 14 (13-15) [6-21] |

ERL, Erlangen; IQR: interquartile range.; n* data available for 101 out of 127 volunteers

### 4. Characteristics of cohorts analyzed in laboratory 2 (Erlangen, Germany)

Healthcare workers were offered analysis of their neutralizing antibody responses to SARS-CoV-2 two to three weeks after the second BNT162b2 mRNA vaccination. Given the uncertainties in the immunogenicity of heterologous prime boost vaccinations, recipients of a priming vaccination with ChAdOx1 nCoV-19, who had received a second vaccination with ChAdOx1 nCoV-19 or BNT162b2 mRNA on April 29th, 2021, at the public vaccination center in Erlangen were offered to register by phone for blood sampling appointments 14 to 16 days after the second vaccination. Blood samples and information on age, sex, and dates of first and second vaccination were obtained from 316 volunteers (for Summary see Table S2). Volunteers reporting prior SARS-CoV-2 infection were excluded.

5. **Supplementary Figure.**

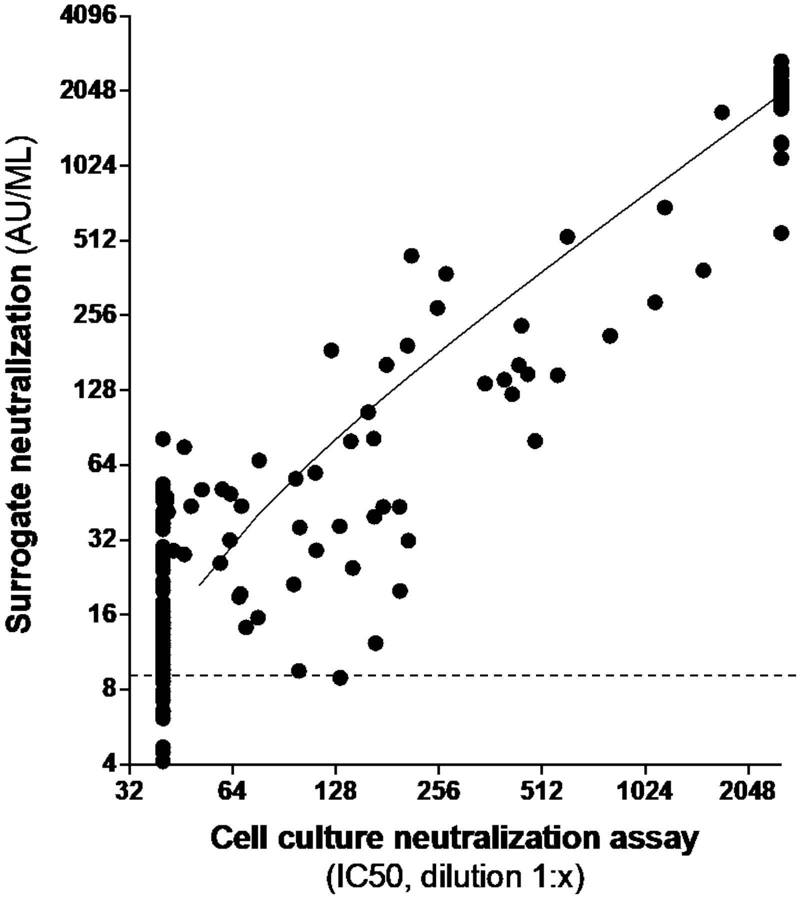


**Figure S1: Correlation of the surrogate neutralizing antibody assay and a cell-culture based SARS-CoV-2 infection inhibition assay.** 220 sera obtained from COVID-19 convalescent or vaccinated donors were analyzed 1) in a cell-culture based assay using SARS-CoV-2 wild-type and 2) in a competitive chemo-luminescence immunoassay (iFlash-2019-nCoV NAb assay, Yhlo, Shenzen, China). The surrogate neutralization assay detects antibodies that are able to compete with binding of a recombinant extracellular domain of the SARS-CoV-2 receptor ACE2 to a recombinant receptor-binding domain of the SARS-CoV-2 S1 protein coated on microparticles and thus indicates the neutralization activity of sera. Activity is determined in arbitrary units (AU). Cut-off is 10 AU/ml, indicated by the dotted line. For the cell-culture assay, Vero E6 cells were infected with SARS-CoV-2 (Wuhan strain, D614G variant) at a multiplicity of infection of 0.06 incubated with test sera in a serial 2-fold dilution form 1:40 to 1:2560. After 24 hours, cells were fixed for an in-cell ELISA using anti-dsRNA J2 antibody (Jena Bioscience, Jena, Germany) to detect SARS-CoV-2 infection. Using the resulting inhibition curve, the inhibitory 50% concentration (IC50) given as dilution 1:x was determined. All positive values obtained are plotted. Analysis revealed a linear regression with a slope of 0.7863 (0.7566 to 0.8160) and an $R$ squared of 0.9258 being significantly non-zero ($p<0.001$).

### 6. Ethics Statement

The retrospective analysis plan on the comparison of the SARS-CoV-2 antibody levels in response to different vaccine regimens was approved by the local ethics committees in Erlangen (Az. 202_21 Bc) and Munich (Az. 26/21 und Az. 330/21 S).


### 7. Funding Statement

Funded by the German Centre for Infection Research (DZIF), the European Union's “Horizon 2020 Research and Innovation Programme" under grant agreement No. 101037867 (VACCELERATE), the “Bayerisches Staatsministerium für Wissenschaft und Kunst” for the CoVaKo-2021 and the For-COVID projects and the Helmholtz Association via the collaborative research program “CoViPa”. Further support was obtained from the Federal Ministry of Education and Science (BMBF) through the “Netzwerk Universitätsmedizin”, project “B-Fast”, and the Bavarian Research Foundation via project “CoV-Rapid”.

### 8. Acknowledgements

We thank Thomas Wochnig, Anna-Marja Jetzlsperger, Philip Hagen, Kirsten Fraedrichfor and Norbert Donhauser, excellent technical assistance. We are grateful to Martina Rupp, Oana Wehrmann and Nives Schwerdtner for their help in arranging the appointments for the vaccinees and setting up the blood sampling site at the Microbiology Institute in Erlangen and to all medical doctoral students for their assistance.
